# A Usability Evaluation of YouDiagnose: Artificial Intelligence Powered Physician Consultation

**DOI:** 10.1101/2022.12.20.22283710

**Authors:** Aswini Misro, Naim Kadoglou, Nishikant Mishra, Paul Whittington, Huseyin Dogan

## Abstract

The COVID-19 Pandemic has resulted in a forced transition to tele-medicine, where history-taking and clinical assessments are performed remotely during video or telephonic consultations. While telemedicine has added to safety and social distancing during the pandemic, the manual and resource-intense process of telephonic and video consultations has not helped to ease the patient backlog, rather has added to this snowballing issue. This paper describes about YouDiagnose pre-consultation exercise that automates patient triage and clinical assessment using artificial intelligence technologies delivered through either a Smart Questionnaire or Chatbot. A usability evaluation was conducted with participants from the Patient and Public Involvement and Engagement Senate (PIES) of the Innovation Agency (an Academic Health Science Network) Qualitative feedback was obtained from the participants on both modalities and quantitative feedback in the form of the System Usability Scale (SUS), comparing the usability of both interaction modalities. The SUS scores were analysed using the Adjective Rating Scale that revealed the Smart Questionnaire had ‘Good Usability’ compared to ‘OK Usability’ of the Chatbot. The results shows the user experience and untapped potential of process automation and artificial intelligence in clinical services.

## 1 Introduction

In 2020, the Health Secretary of the United Kingdom (UK) Matt Hancock said the following in light of the COVID-19 pandemic, “We have moved to a principle of digital first in primary care and with outpatients: unless there are clinical or practical reasons, all consultations should be done by telemedicine” [1]. While the pandemic has mandated the introduction of such drastic measures, the evidence about patient outcomes, cost-effectiveness, safety, technical issues, quality of consultations and the overall impact, is usually mixed and based on reports from small studies. The randomised control trials conducted on video consultations in hospital outpatient clinics mainly focused on managing chronic long-term conditions [5,6,7,8]. Most of these studies have reported a high degree of patient and clinician satisfaction with no difference in disease progression, compared with the traditional face-to-face consultation mode. Remote consultations are often seen to be lacking information compared to face-to-face consultations and can have many challenges [9,10]. Furthermore, there is a significant divide between physicians concerning their preference for video or telephone consultations [12,13].

## 2 Related Work

We have conducted Literature Reviews into the current challenges of video consultations. Even though they can provide a number of benefits, such as increased convenience and reduced infection transmission, video consultations have the disadvantage of being time-limited. According to research carried out and published in the British Medical Journal, General Practitioners (GPs) significantly asked a smaller number of questions concerning presenting symptoms with low adherence to guidelines, less thorough clinical examination and less advice on lifestyle [20]. Moreover, a recent study conducted in 2019-2020 on general practice in the UK showed that digital-first access models using online, telephone or video consultations are likely to increase the GP’s workload by 25%, 3%, and 31%, respectively [21]. Our review of sources [9,10,11,12,13,14,15,16,17,18] highlighted a number of challenges of remote video/telephone consultations, including slow internet speed, poor audio/video quality, security and privacy concerns and patients preferring face-to-face consultations.

We identified the three main challenges as being:

- Technical issues and unfamiliarity with technology resulting in a loss of information during the remote consultation. Furthermore, the diagnosis of cancer can involve physical examination that is impossible to perform remotely.
- Studies show that people over the age of 60 years are less likely to use remote video consultations as a primary mode.
- GP consultations are often limited to 10 minutes, which is often an insufficient duration for a detailed consultation.

The impact of complete virtualisation of patient consultation has not fully been investigated. However, there have been examples of lapses and delays in diagnosis of patients. The COVID-19 pandemic has been said to be one of the contributing factors of low cancer referrals [19]. This could also be caused by missed opportunities in the early diagnosis during telephone or video consultations, due to reduced examination findings. Additionally, remote consultations can be less suitable for vulnerable patients from low socioeconomic backgrounds, compared to patients from high socioeconomic settings, which increases the inequalities surrounding early cancer diagnosis.

As a result of virtualisation of clinical consultations, we have been carrying out research and development work into hybrid clinical consultations. In many clinical consultations, the patient is asked to complete a pre-consultation questionnaire, where the structure of the questionnaire does not allow for any customisation or personalisation that may be required for a particular circumstance or patient. This can lead to reduced patient satisfaction due to the questionnaire being time consuming to complete.

The four main issues associated with the structured questionnaire for inpatient services are:

1. Structured questionnaires are not always relevant to certain situations. Therefore, the patient may not be 100% truthful in their answer to mandatory questions and can ignore questions that are non-mandatory.
2. Questions can be difficult to interpret for patients, as there is no explanation provided and this can lead to reduced user comprehension.
3. Multiple choice questions may not fully capture a patient’s complete diagnosis, as they often consist of closed questions, with a set number of options. These may not be applicable to the complete diagnosis of the patient or consider the emotional aspect.
4. Completing long questionnaires with repetitive questions can lead to patient fatigue, becoming less inclined to engage in the process.

Prior to the development of the YouDiagnose Chatbot and Smart Questionnaire, we conducted interviews with healthcare professionals (n=6), who work in the National Health Service (NHS) secondary care. These interviews focused on the challenges encountered due to the COVID-19 pandemic and the impact as a result on patient consultation. One participant highlighted, “*as it takes longer to consult a patient over the phone, fewer patients used to be booked during the initial days of the pandemic; however, in the last few months, the number has gone up to the pre-pandemic level*”. It was evident that some healthcare professionals do not have access to video consultations and had to rely on telephone consultations with patients. An additional challenge was not being able to examine a patient, resulting in a reduction of their confidence when diagnosing patients. However, it was acknowledged that telephone consultations were necessary for certain situations.

It is anticipated that these challenges can be reduced through the adoption of artificial intelligence and chatbot technology to provide personalised patient questionnaires.

## 3 YouDiagnose Chatbot and Smart Questionnaire

Our solution is to use the YouDiagnose Chatbot and Smart Questionnaire to provide a pre-consultation through artificial intelligence-led technologies. This was applied to carry out breast cancer risk assessments, by asking patients specific questions regarding their various cancer risk factors, such as habits and physical body characteristics.

In the Smart Questionnaire patients complete a series of consecutive questions about their diagnosis. Using the Chatbot, chat messages are sent to the user in the form of questions that provide a set of answers and the user selects the relevant answer though the touchscreen.

Both of these techniques use a smart recognition pattern, whereby the user’s response to a question determines the next set of questions. An example is when a patient has undergone a breast biopsy and the Smart Questionnaire or Chatbot ask questions relating to the details of the breast biopsy, therefore producing a personalised questionnaire. This type of questionnaire is more efficient to complete, as it is tailored to the patient’s specific diagnosis. As the diagnosis process is less time consuming, this has the potential to increase patient satisfaction.

### 3.1 Usability Evaluation

The usability evaluation of YouDiagnose Chatbot and Smart Questionnaire was conducted in association with the Patient and Public Involvement and Engagement Senate (PIES) of Innovation Agency and the Academic Health Science Network (AHSN) for the North West Coast. The Patient and Public Involvement and Engagement Senate operates in Lancashire, South Cumbria, Cheshire and Merseyside. They provide a unique perspective to various healthcare projects, improving the patient experience for different healthcare technologies across the northwest coast, including the Clatterbridge Cancer Centre in Liverpool. The North West Coast AHSN is one of 15 AHSNs in the United Kingdom that collectively forms the innovation section of the NHS.

To evaluate the usability of the Chatbot and Smart Questionnaire interaction modalities, a study was conducted involving PIES Senate members, cancer carers and survivors. A usability study pack was designed by the Digital Health department of Bournemouth University at the outset. This usability pack contained various types of survey questions the user had to respond after they has finished their interaction with the technology.

The Senate members were introduced through a virtual PIES meeting, where YouDiagnose delivered a presentation on the two interaction modalities of the Chat-bot and Smart Questionnaire. The attendees were then sent the survey by email and instructed to use YouDiagnose and provide feedback. Social media was used to contact a number of cancer carers and survivors, who were also invited to conduct the evaluation. Out of the 61 participants who were contacted for the study, 39 responded and agreed to conduct the evaluation. The 39 participants completed the evaluation of both modalities and their demographics are shown in Table 1. All participants owned a smartphone and had broadband internet access at home.

**Table 1:** Usability Evaluation Participants Demographics.

| Demographic | Participant Numbers |
| --- | --- |
| Age | 18 – 25: 5 |
|  | 26 – 35: 6<br>36 – 49: 6<br>50 – 65: 6<br>Over 65: 16 |
| **Gender** | Male: 18<br>Female: 20<br>Non-binary: 1 |
| **Ethnicity** | Asian: 12<br>Black African, Caribbean or Black British: 3<br>British White: 18<br>Mixed or Multiple Ethnic Group: 3<br>Other Ethnic Group: 3 |
| **Primary Language** | English: 18<br>European Foreign Languages: 3<br>Other Foreign Languages: 18 |
| **Highest Education Level** | College Graduate: 19<br>High School Graduate: 6<br>Postgraduate Studies: 3<br>Professional, e.g. PhD: 11 |
| **Household Income (£)** | Less than 15,000: 4<br>15,000 to 24,999: 5<br>25,000 to 44,999: 20<br>45,000 to 64,999: 9<br>65,000 to 99,999: 1 |

Each participant was provided with a user information pack consisting of a link to the YouDiagnose web portal that provided an overview of the technologies, including videos and text descriptions. The pack also contained links to test the Chatbot and Smart Questionnaire and a survey where participants could express their experience and views of the modalities. This included the use of the System Usability Scale (SUS), to provide quantitative feedback for both modalities and an accurate assessment of the usability. The advantage of SUS is that the technique produces a single usability score that can be interpreted using the Adjective Rating Scale. This scale determines a usability rating, between ‘Worst Imaginable’ and ‘Best Imaginable’, as described in Table 2. The participants who did not respond to the initial contact, were sent three follow-up reminders to complete the evaluation, through telephone, email and social media messages.

**Table 2:**
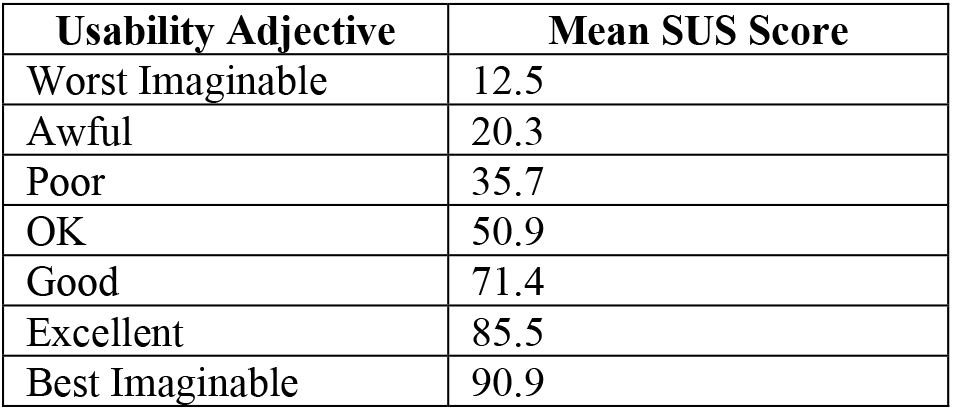
Adjective Rating Scale Thresholds.

### 3.2 Qualitative Feedback

The evaluation participants provided qualitative feedback for both the Smart Questionnaire and the Chatbot through a survey. Both modalities received positive usability feedback, as well as suggestions for improvements.

Participants found the Smart Questionnaire easy to use with language that was simple to understand, although there were suggestions that further graphics could be included. It was stated that the Smart Questionnaire could provide links to further information, as well as details of how to calculate alcohol units so that they could provide an accurate representation of their lifestyle. There were some technical issues raised, including rotating the mobile device causing display issues with the Smart Questionnaire. It was highlighted that further audio/visual features could be included, for example, the ability to read text aloud. It was also suggested that a Frequently Asked Questions section would be a useful addition, in providing answers to common user questions. The Chatbot was found to be easy to use with an interface that was simplistic, with the technology saving time during the diagnosis process. However, a participant thought that further background information could be provided by the Chatbot, including hyperlinks to further information. When using both modalities, some participants encountered loss of data when the device had a technical failure, e.g. loss of battery charge. One participant highlighted that the questionnaire they completed, contained a great number of questions which they found time consuming and cumbersome to complete.

### 3.3 System Usability Scale Results

The SUS Questionnaires completed by the participants from both the Smart Questionnaire and the Chatbot were analysed using the Adjective Rating Scale, to calculate the mean usability score between 0 and 100. This score can be interpreted to determine the level of usability between ‘Worst Imaginable’ and ‘Best Imaginable’, with the thresholds stated in Table 2.

Using the survey, the participants rated their strength of agreement for questions relating to the usability of each modality. These questions included, “I would like to use the Smart Questionnaire to inform my diagnosis”, “I think I would need support to use the Chatbot” and “I think most people would learn to use the Smart Questionnaire very quickly”. The SUS scores were calculated for each participant and an average SUS score was interpreted using the Adjective Rating Scale. The Smart Questionnaire achieved a SUS score of 78.1, indicating “Good Usability”. This supported the qualitative feedback where participants highlighted that the user interface was easy to use and the language was simplistic. The Chatbot achieved a SUS score of 71.3 and was therefore interpreted as “OK Usability”. The reduction in the usability of the Chatbot could have been as a result of the modality having an insufficient amount of background information available.

## 4 Discussion

This study compared the usability of the YouDiagnose Chatbot and Smart Questionnaire that uses artificial intelligence to conduct breast cancer risk assessments based on a number of factors relating to the patient’s lifestyle, e.g. exercise and alcohol consumption habits. The two modalities enable patients to interact with the assessment process, either through completing a Smart Questionnaire that adapts to a patient’s diagnosis or through interaction with a Chatbot. The usability of both of these interaction modalities was assessed using the System Usability Scale, where SUS scores were interpreted using the Adjective Rating Scale. The SUS scores for the Chatbot and Smart Questionnaire revealed, ‘Good Usability’ and ‘OK Usability’ respectively, indicating that the Smart Questionnaire was the more efficient interaction modality. Further assessments of the usability were obtained through qualitative feedback of the user experience during interaction. This highlighted the possible causes of the decreased usability, in that some participants found the Chatbot to contain an excessive number of questions that were time-consuming to complete. Additionally, the usability could be further increased through the provision of information surrounding the patient’s diagnosis and answers to Frequently Asked Questions.

The evaluations identified that the interface of both modalities were easy to use and simple to understand. However, there were some technical deficiencies with both modalities in that the web-based version was not supported by certain browsers and the application did not function correctly when the orientation of the smartphone was changed. Overall, the participants emphasised that YouDiagnose was an effective breast cancer risk assessment method, through the use of artificial intelligence technologies for patient diagnosis.

## 5 Conclusions and Future Work

The participants’ feedback from the usability evaluations will be addressed by future development of the Chatbot and Smart Questionnaire. This will include the provision of further information surrounding the patient’s diagnosis and resolution of the technical defects surrounding device orientation and web browser compatibility. A further evaluation study will then be conducted to reassess usability, using SUS and comparing the updated SUS scores using the Adjective Rating Scale. In addition to SUS, NASA Task Load Index (TLX) [23] will be applied to measure the users’ workload demands when using the two modalities. NASA TLX measures Physical, Mental, Temporal, Performance, Effort and Frustration demands and is highly accessible in a range of freely available formats, including an iOS application and a printed questionnaire [24].

The YouDiagnose Chatbot and Smart Questionnaire provides a solution for healthcare professionals, to determine the risk of breast cancer remotely, through artificial intelligence technology. The Smart Questionnaire was considered to have the highest usability score, but the Chatbot and Smart Questionnaire were both shown to be suitable methods to initially determine breast cancer risk, prior to patients having a consultation. The need for such a diagnosis method has increased due to the COVID-19 pandemic, with a greater reliance on remote consultations. It is evident through the usability evaluation results, that both the YouDiagnose Chatbot and Smart Questionnaire modalities provide patients with an effective breast cancer risk assessment.

## Data Availability

All data (not related to IP) produced in the present study are available upon reasonable request to the authors.

## Notes

### Competing Interest Statement

The authors have declared no competing interest.

### Funding Statement

This study did not receive any funding

### Author Declarations

Ethics committee of YouDiagnose Limited gave ethical approval for this work.

## References

1. COVID-19: video consultations and homeworking, https://www.bma.org.uk/advice-and-support/covid-19/adapting-to-covid/covid-19-video-consultations-and-homeworking, last accessed 2021/06/23.

5. Freeman, K.A., Duke, D.C., Harris M.A.: Behavioral health care for adolescents with poorly controlled diabetes via Skype: does working alliance remain intact? J. Diabetes Sci. Technol. (7), 727–35 (2013).

6. Hansen, C.R., Perrild, H., Koefoed, B.G., Zander, M.: Video consultations as add-on to standard care among patients with type 2 diabetes not responding to standard regimens: a randomised controlled trial. Eur. J. Endocrinol. (176), 727–36 (2017).

7. Harris, M.A., Freeman, K.A., Duke, D.C.: Seeing is believing: using Skype to improve diabetes outcomes in youth. Diabetes Care (38), 1427–34 (2015).

8. Real-time (synchronous) telehealth in primary care: systematic review of systematic reviews Canadian Agency for Drugs and Technologies in Health 2008. https://www.crd.york.ac.uk/CRDWeb/ShowRecord.asp?AccessionNumber=32008000106&AccessionNumber=32008000106, last accessed 2021/06/23.

9. Donaghy, E., Atherton, H., Hammersley, V., McNeilly, H., Bikker, A., Robbins, L., Campbell, J., McKinstry, B: Acceptability, benefits, and challenges of video consulting: a qualitative study in primary care. British J. Gen. Pract., 69, 586–9 (2019).

10. Hammersley, V., Donaghy, E., Parker, R., McNeilly, H., Atherton, H., Bikker, A., Campbell J., McKinstry B.: Comparing the content and quality of video, telephone, and face-to-face consultations: a non-randomised, quasi-experimental, exploratory study in UK primary care. British J. Gen. Pract., 69(686) 595–604 (2019).

11. What will new technology mean for the NHS and its patients? Four big technological trends, https://www.kingsfund.org.uk/sites/default/files/2018-06/NHS_at_70_what_will_new_technology_mean_for_the_NHS_0.pdf, last accessed 2021/06/23.

12. National Institute for Health Research: Alternatives to face-to-face consultation with a GP. GP Primary Care Digest (2017).

13. Fatehi, F., Menon, A., Bird, D.: Diabetes care in the digital era: a synoptic overview. Curr. Diab. Rep., 18, 38 (2018).

14. The NHS long term plan, https://www.longtermplan.nhs.uk/wp-content/uploads/2019/01/nhs-long-term-plan-june-2019.pdf, last accessed 2021/06/23.

15. Zanaboni, P., Wootton, R.: Adoption of routine telemedicine in Norwegian hospitals: progress over 5 years. BMC Health Serv. Res., 16, 496 (2016).

16. Greenhalgh, T., Shaw, S., Wherton, J., Vijayaraghavan, S., Morris, J., Bhattacharya, S., Hanson, P., Campbell-Richards, D., Ramoutar, S., Collard, A., Hodkinson, I.: Real-world implementation of video outpatient consultations at macro, meso, and micro levels: mixed-method study. J. Med. Internet. Res., 20, e150 (2018).

17. Safe, seamless and secure: evolving health and care to meet the needs of modern Australia, https://conversation.digitalhealth.gov.au/australias-national-digital-health-strategy, last accessed 2021/06/23.

18. Appointments in general practice England, https://digital.nhs.uk/data-andinformation/publications/statistical/appointments-in-general-practice/august-2020, last accessed 2021/06/23.

19. McCall, B.: Could telemedicine solve the cancer backlog? The Lancet Digital Health, 2(9), 456–57 (2020).

20. Tsiga, E., Panagopoulou, E., Sevdalis, N., Montgomery, A., Benos, A.: The influence of time pressure on adherence to guidelines in primary care: an experimental study. BMJ Open, 3, e002700 (2013).

21. Salisbury, C., Murphy, M., Duncan, P.: The Impact of Digital-First Consultations on Workload in General Practice: Modeling Study. J. Med. Internet Res., 22(6), e18203 (2020).

22. NASA TLX: Task Load Index, https://humansystems.arc.nasa.gov/groups/tlx/, last accessed 2021/07/28.

23. Everything You Need to Know About the NASA Task Load Index (NASA-TLX), https://research-collective.com/nasa-tlx/, last accessed 2021/07/28.

